# Pollen explains seasonality of flu-like illnesses including COVID-19

**DOI:** 10.1101/2020.06.05.20123133

**Authors:** Martijn J. Hoogeveen, Eric C.M. van Gorp, Ellen K. Hoogeveen

## Abstract

Current models for flu-like epidemics insufficiently explain multi-cycle seasonality. Meteorological factors alone, including the associated behavior, do not predict seasonality, given substantial climate differences between countries that are subject to flu-like epidemics or COVID-19. Pollen is documented to be allergenic, it plays a role in immuno-activation and defense against respiratory viruses, and seems to create a bio-aerosol that lowers the reproduction number of flu-like viruses. Therefore, we hypothesize that pollen may explain the seasonality of flu-like epidemics, including COVID-19, in combination with meteorological variables.

We have tested the Pollen-Flu Seasonality Theory for 2016-2020 flu-like seasons, including COVID-19, in the Netherlands, with its 17.4 million inhabitants. We combined changes in flu-like incidence per 100K/Dutch residents (code: ILI) with pollen concentrations and meteorological data. Finally, a predictive model was tested using pollen and meteorological threshold values, inversely correlated to flu-like incidence.

We found a highly significant inverse correlation of r(224)= −0.41 (p < 0.001) between pollen and changes in flu-like incidence, corrected for the incubation period. The correlation was stronger after taking into account the incubation time. We found that our predictive model has the highest inverse correlation with changes in flu-like incidence of r(222) = −0.48 (p < 0.001) when average thresholds of 610 total pollen grains/m^3^, 120 allergenic pollen grains/m^3^, and a solar radiation of 510 J/cm^2^ are passed. The passing of at least the pollen thresholds, preludes the beginning and end of flu-like seasons. Solar radiation is a co-inhibitor of flu-like incidence, while temperature makes no difference. However, higher relative humidity increases with flu-like incidence.

We conclude that pollen is a predictor of the inverse seasonality of flu-like epidemics, including COVID-19, and that solar radiation is a co-inhibitor.

## 1. Introduction

Current models for flu-like epidemics insufficiently explain multi-cycle seasonality. Meteorological factors alone do not fully explain the seasonality of flu-like epidemics (Tamerius et al., 2011) or COVID-19 (Yao et al, 2020). Pollen is documented to be allergenic (Klemens et al, 2007; Rosenwasser, 2011; Howarth, 2000), and it plays a role in immuno-activation (Brandelius et al, 2020. Furthermore, allergic diseases are absent as a comorbid condition of COVID-19 (Zhang et al., 2020; Dong et al., 2020). Explaining this, Licari et al. (2020) found that allergic children have a significantly higher eosinophil count than COVID-19 patients, whereby eosinophils are known to clear viral load, and contribute to the recovery from viral infections, supposedly including COVID-19 (Lindsley et al, 2020). A further explanation is provided by Jackson et al. (2020), who proved that allergic sensitization and allergen natural exposure are inversely related to ACE-2 expression, whereby it is known that COVID-19 uses the ACE-2 receptor to enter the host (Wan et al., 2020). Finally, histamine and IgE serum levels are elevated in allergic rhinitis and asthma patients – an allergenic disease that is also underreported as comorbid condition for COVID-19 – which downplay certain other anti-viral responses (plasmacytoid dendritic cells and interferon-α) but might thus prevent the cytokine storm and hyper-inflammation that typically mark severe respiratory diseases, including COVID-19 (Carli et al., 2020). Similarly, it is reported that pollen might suppress interferon-λ and pro-inflammatory chemokine responses in non-allergic subjects and it is found to be correlated to rhinovirus-positive cases (Gilles et al, 2020), but not to other flu-like virus-positive cases (Nivel.nl, 2020). It is hypothesized that exposure to and immunization for such ordinary cold viruses, especially corona cold viruses, could provide some protection against COVID-19, in the same way that the uptake of the common influenza vaccine seems to be inversely correlated to COVID-19 deaths (Marín-Hernández et al., 2020).

Recently, we identified pollen bio-aerosol as a discrete seasonal factor in inhibiting flu-like epidemics during the period 2016 to 2019 in the Netherlands (Hoogeveen, 2020). In this epidemiological study, we found strong inverse correlations between allergenic pollen concentrations and hay fever on the one hand, and flu-like incidence on the other. The study was based on the persistent observation that the pollen and flu season predictably alternate each other in moderate climate zones, and the absence of sufficient meteorological explanations (Tamerius et al., 2011). We further observed that the passing of pollen threshold values of around 100 allergenic pollen grains/m^3^, reliably mark the onset and decline of moderate flu-like epidemic lifecycles, and thus might be used as predictor. Such a concentration of allergenic pollen makes sense, as in an overview of real-life studies, clinical pollen threshold values are observed between 1 and 400 grains/m^3^, whereby the first symptoms are typically observed in the range of 1 – 50 grains/m^3^ (De Weger et al, 2013), depending on the country, period and vegetation, and, probably, the susceptibility of the subjects to allergens.

The seasonality of respiratory viral infections has been recognized for thousands of years in temperate regions (Moriyama et al., 2020). Seen in more in detail, virologists observed that cold, and flu-like epidemics (e.g., influenza and corona-caused) “go away in May” in the Northern Hemisphere, while emerging in the Southern Hemisphere with its opposite seasonality, only to re-emerge in the Northern Hemisphere during its next autumn and winter, but in a slightly mutated form. Furthermore, all new flu-like pandemics since 1889 typically emerged in the Northern Hemisphere at the tail-end of respective flu-seasons (Fox et al., 2017). Clearly, the current COVID-19 pandemic is no exception. The emergence of COVID-19 and other pandemics at the tail-end of the flu season makes sense. It takes time for a spontaneous new crossover virus with a sufficiently high reproduction number (Ro) – for SARS-CoV-2 it is estimated to be initially around 3 (Liu et al., 2020) – to develop from patient 0 to become a fully-fledged pandemic during the flu season in the Northern Hemisphere. The Northern Hemisphere, with its larger and denser populations, is more likely to be the initial breeding ground for a new flu-like pandemic than the Southern Hemisphere. Furthermore, Fox et al. showed that most flu-like pandemics are multi-wave, whereby the initial wave at the tail-end of flu season is typically short-lived. This gives rise to the idea that COVID-19 is also subject to such multi-wave seasonality (Kissler et al., 2020), because the distribution of community outbreaks is consistent with the behavior of seasonal respiratory viruses (Sajadi et al., 2020), and has a short wave at the tail end of the 2019/2020 flu-like season in the Northern Hemisphere in the temperate climate zone.

Numerous studies try to explain flu-like seasonality with meteorological factors such as sunlight, including UV radiation (Schuit et al., 2020), temperature and humidity (Chong et al., 2020; Shaman et al, 2011). However, Postnikov (2016) concluded that ambient temperature is not a good predictor for influenza seasonality in the Netherlands, and inconsistent correlations also exist for the relationships between COVID-19 and temperature (Toseupu et al, 2020; Xie & Zhu, 2020; Ma et al 2020; Qi et al, 2020). Furthermore, findings about the relationships between humidity and influenza (Soebiyanto et al., 2014), and humidity and COVID-19 (Ahmadi et al, 2020; Ma et al, 2020; Qi et al, 2020) are equally inconsistent. Although UV light is detrimental for the flu-like virus aerosol under laboratory conditions, associated with immuno-activation (Abhimanyu & Coussens, 2017; Tan & Ruegiger, 2020) and circadian rhythms regulating lung immunity (Nosal et al., 2020), the onset of the flu season, from mid-August in the Netherlands, coincides with an annual peak in hot, sunny days and is still in the middle of the summer season. According to Yao et al. (2020), for a decrease of COVID-19 infections neither high UV values nor high temperatures are good predictors. The contradictory findings related to COVID-19, understandably based on the analysis of a limited part of the year and disease cycle, might be partly due to sub-seasonal bias and unstandardized data-collection methods. By sub-seasonal bias we mean that if only a part of a season or cycle is analyzed, overly specialized conclusions can be drawn that cannot be generalized to the whole season or cycle.

Nevertheless, these meteorological variables are known factors in flowering and pollen maturation and dispersion. Meteorological variables, such as increased solar radiation and temperature – among others the absence of frost – not only trigger flowering and pollen maturation, they also affect the pollen bio-aerosol formation: dry and warm conditions stimulate pollen to become airborne. Rain, in contrast, makes pollen less airborne, and cools the bio-aerosol down. Very high humidity levels (RH 98%) are even detrimental to pollen (Guarnieri, 2006). An RH 98% effect on pollen could thus provide an alternative explanation as to why flu-like incidence in tropical countries is higher during the rainy season, and reduced during the rest of the year.

We hypothesize that pollen bio-aerosol has an inverse effect on flu-like incidence, including COVID-19 (see Figure 1), whereby pollen is known to be triggered and influenced by meteorological variables, which can then jointly explain the seasonality of flu-like incidence. This indirect explanation of the pollen effect is based on the fact that pollen bio-aerosol and UV light exposure lead to immuno-activation, and sometimes allergic symptoms, which seem to protect against flu-like viruses, or at least severe outcomes from them. The indirect pollen effect is explained by the spread of pollen bio-aerosol under sunny and dry conditions, which *might* absorb viral bio-aerosol, and *might* also become an agent that leads to an increased exposure of flu-like viruses to virus-degrading solar radiation, including the UV spectrum, and – possibly – anti-viral phytochemicals in pollen.

**Figure 1:**
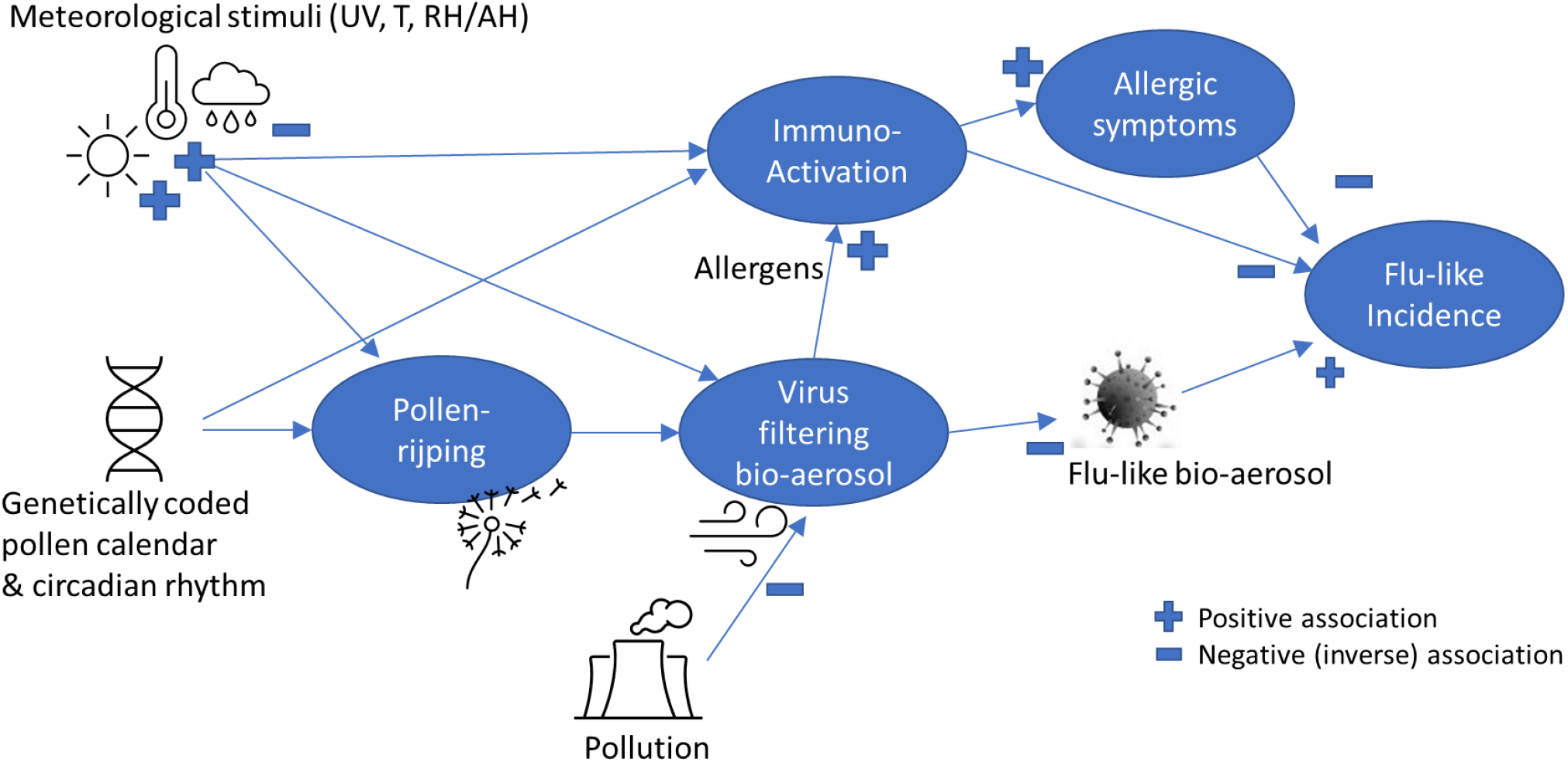
A simplified theoretic model explaining pollen-flu inverse seasonality, whereby pollen might have an allergenic (I) and/or immune-triggering (II) function, and/or a direct (III) “antiviral” effect on flu-like bio-aerosol, inhibiting flu-like epidemic incidence in combination with meteorological conditions and triggers.

To further understand the impact of pollen as an environmental factor influencing the life cycle of flu-like epidemics, the objective of this study is to determine the correlations of pollen and meteorological variables with (changes in) flu-like incidence and develop and test a discrete predictive model that combines pollen and meteorological co-inhibitors. Our main hypothesis, therefore, is that pollen is the missing link, jointly explaining with certain meteorological variables, flu-like seasonality, and that a compound threshold based factor – combining detected flu-inhibitors – is a good unified predictor of such seasonality. Regarding COVID-19, we have limited ourselves to observing whether or not COVID-19 at the tail-end of the 2019/2020 flu-like season is able to break with the flu-like seasonality pattern.

## 2. Methods

To study the relationship between pollen and flu-like incidence in the Netherlands, we used the public datasets of Elkerliek Hospital (Elkerliek.nl) about the weekly allergenic, low-level allergenic and total pollen concentrations in the Netherlands in grains/m^3^, whereby for 42 types of pollen particles the numbers are counted and averaged per day per 1m^3^ of air. The common Burkard spore trap was used, through which a controlled amount of air was ingested. The applied classification and analysis method conforms to the EACCI (European Academy of Allergology and Clinical Immunology) and the EAN (European Allergy Network) standards. Allergenic pollen includes nine types of particles that are classified as moderate (*Corylus, Alnus, Rumex, Plantago* and *Cedrus Libani*), strong (*Betula* and *Artemisia*), or very strong allergenic (*Poaceae* and *Ambrosia*). Additionally, we included low-level allergenic pollen concentrations in addition to the allergenic ones because we assume that they will also have effects. Low-level allergenic pollen includes the other 33 particles that are classified as non-allergenic to low-level allergenic (*Cupressaceae, Ulmus, Populus, Fraxinus, Salix, Carpinus, Hippophae, Fagus, Quercus, Aesculus, Juglans, Acer, Platanus, Pinus, Ilex, Sambucus, Tilia, Ligustrum, Juncaceae, Cyperaceae, Ericaceae, Rosaceae, Asteraceae, Ranunculaceae, Apiaceae, Brassicaceae, Urtica, Chenopodiaceae, Fabaceae, Humulus, Filipendula*, and *Indet*). Total pollen concentration is the sum of the average allergenic and low-level allergenic pollen concentrations. Advantages of using the total pollen metric are that there are hardly any 0 values (only 3 out of 266), and we did not need to limit ourselves to just parts of the seasonal cycle, which might introduce sub-seasonal bias into our research. We also assumed that long-distance pollen transport is accounted for, as foreign pollen will also be counted by a pollen measuring station that works all year round.

Furthermore, we used the data from the Dutch State Institute for Public Health (RIVM.nl) gathered by Nivel (Nivel.nl) about weekly flu-like incidence (WHO code “ILI” - Influenza Like Illnesses) reports at primary medical care level, per 100,000 citizens in the Netherlands. Primary medical care is the day-to-day, first-line healthcare given by local healthcare practitioners to their registered clients as typical for the Netherlands, with its current population of 17.4 million. The reports relate to a positive RIVM laboratory test for ILI after a medical practitioner diagnosed ILI after a consultation, whether that leads to hospitalization or not. The ILI metric is according to a standardized WHO method, given that ILI data is gathered and compared globally. ILI is defined by the WHO as a combination of a measured fever of ≥ 38°C, and a cough, with an onset within the last 10 days. The flu-like incidence metric is a weekly average based on a representative group of 40 primary care units, and calculated using the number of influenza-like reports per primary care unit divided by the number of patients registered at that unit. This is then averaged for all primary care units and then extrapolated to the complete population. The datasets run from week 1 of 2016 up to week 18 of 2020 (n = 226 data points) to include the recent COVID-19 pandemic at the tail-end of the 2019/2020 flu-like season. To underpin the relative importance of COVID-19: SARS-CoV-2 has been detected in the Netherlands since week 9, 2020. According to the figures of Nivel.nl (2020; see Figure 2), from week 13 onwards SARS-CoV-2 is the outcome of the (vast) majority of positive tests for patients at primary care level with flu-like complaints, and by week 18 100% of positive tests indicate SARS-CoV-2 (other tested viruses are five Influenza A and B subtypes, RSV, Rhinovirus and Enterovirus).

**Figure 2:**
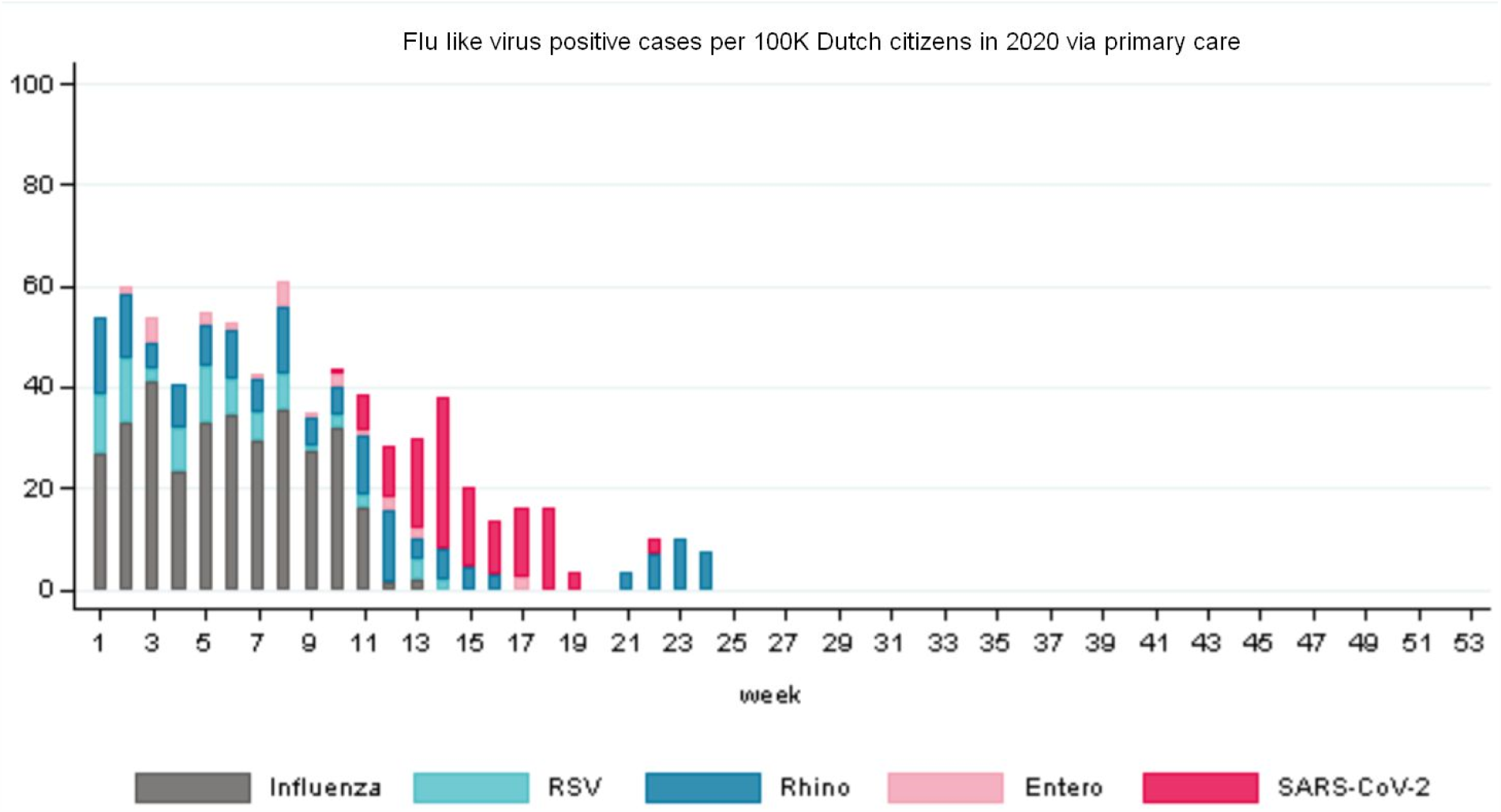
The overview of flu-like virus positive cases in 2020 (till week 24) shows that from week 13 onwards SARS-CoV-2 is dominant. The Rhinovirus cases spike is outside the timeline of our research (source: Nivel.nl, 2020).

Furthermore, we also included meteorological datasets from the Royal Dutch Meteorological Institute (KNMI.nl), including average relative humidity/day, average temperature/day and global solar radiation in J/cm^2^ per day as an indicator of UV radiation. These datasets were obtained from the KNMI’s centrally located De Bilt weather station. Next, we calculated the weekly averages for the same periods that featured in the other datasets. De Bilt is traditionally chosen as it provides an approximation of modal meteorological parameters in the Netherlands, which is a small country. Furthermore, all major population centers in the Netherlands, which account for around 70% of the total Dutch population, are within a radius of only 60 kilometers from De Bilt. We therefore assumed in this study that the measurements from De Bilt are sufficiently representative for the meteorological conditions typically experienced by the Dutch population.

To test allergenic versus low-level allergenic pollen assumptions, against hay fever and pre-COVID-19 flu-like incidence, we made use of the hay fever index. The hay fever index is defined as the turnover for hay fever medication, as reported by all Dutch pharmacies to the Dutch Central Bureau of Statistics (CBS.nl) and based on respective ATC codes (especially R01A/R01AC). We used a dataset from week 1 of 2016 up to week 10 of 2019 (n=166 data points), because no further data was made available. For the interpretation of our findings, we assumed for the Netherlands a prevalence of allergic rhinitis that is more or less similar to that in Western Europe, being around 23%, and frequently undiagnosed (Bauchau & Durham, 2004). Furthermore, it can be noted that the prevalence of allergic diseases in general in the Netherlands is around 52% (Van de Ven et al, 2006).

Datasets were complete, except that three weekly pollen concentration measurements were missing (1.3%). This was due to a malfunctioning monitoring station during week 26 of 2016, week 21 of 2017 and week 22 of 2019. These missing measurements appeared to be completely random. We imputed missing values to avoid bias and maintain power. We used a four-week surrounding average to estimate the three missing data points and thus avoid breaking lines in visuals. We checked that the missing data has no material impact on the results by comparing these averages with the data of previous years for similar periods, and by observing whether removal from statistical tests had any effect on outcomes and conclusions.

Regarding the incidence of flu-like symptoms, we calculated the weekly change compared to the previous week (ΔILI=ILI_t_ – ILI_t-1_). This was to obtain an indication of the flu-like epidemic life cycle progression, whereby a decline is interpreted as Ro<1 and an increase as Ro>1 (Ro is the reproduction number of flu-like viruses). Furthermore, to cater, in one time-series metric, for changes in flu-like incidence as well as for an incubation period of up to two weeks, we calculated a three-week moving average (3WMA) of changes in flu-like incidence, of which two weeks are forward looking:

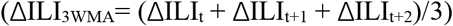

It should be noted that whenever we use the term incubation time, we also mean to include reporting delay (estimated to be around 4.5 days). We have not assumed delay effects for meteorological variables or pollen concentrations, so we have not calculated moving averages for other time series.

Compared with our previous study (Hoogeveen, 2020), there is an overlap in datasets of less than 10%. The datasets are extended by the extension in time, the addition of meteorological datasets and non-allergenic pollen, and the introduction of newly calculated variables, such as total pollen concentration, ΔILI_3WMA_, the compound predictor and the log10 transformations on pollen, ILI and the hay fever index.

We formulated the following statistical null hypotheses for falsification.

H1_0_: there are no inverse correlations for total pollen concentrations with flu-like incidence (corrected for incubation period).

H2_0_: there are no inverse correlations between pollen and *changes* in flu-like incidence (ΔILI or corrected for incubation time: ΔILI_3WMA_).

H3_0_: there is no predictive significance of a discrete model’s compound value, based on thresholds for pollen and meteorological co-inhibitors, related to changes in flu-like incidence (ΔILI_3WMA_).

To understand the role of meteorological variables, to check whether – in our datasets – meteorological variables show their well-established effects on pollen as assumed, and to select co-inhibitors:

H4_0_: meteorological variables – solar radiation, temperature and relative humidity – have no effect on pollen and/or flu-like incidence change (ΔILI_3WMA_).

Low-level allergenic pollen is sometimes known to have a slight allergenic effect. To understand how to interpret adding none-to-low-level allergenic pollen to the total pollen metric, we wanted to verify their effects on the hay fever index:

H5_0_: low-level allergenic pollen has no effect on hay fever and (changes in) flu-like incidence.

Note that with the exception of H5, all hypotheses are related to potential causality: the temporal sequentiality (temporality) of the respective independent variables, and flu-like incidence corrected for incubation period. Whenever we refer to temporality, we mean to indicate that the datasets behave *as if* there is causality, on the understanding that statistics alone cannot prove causality in uncontrolled settings.

### Statistical analyses

Variables are presented with their means (M) and standard deviations (SD).

We calculated correlation coefficients to test the hypotheses and to assess the strength and direction of relationships. As a sensitivity analysis, we also calculated the bootstrapped correlation coefficients. We used the full datasets, to avoid sub-seasonal bias, and by extending the number of years the distortions by incidental and uncontrolled events are supposed to be minimized. However, as a second sensitivity analysis, we removed from the datasets the autumn weeks between 42 and 50, which typically show low pollen concentrations of up to 20 grains/m^3^, which are applied to analyze the main outcome (H2_0_).

Next, linear regression (F-test) on identified inhibitors and interactions was used *descriptively* to determine whether the relationship can (statistically) be described as linear, and to determine the equation using estimates and intercept values, and produce probability, significance level, F-value, and the Multiple R squared correlation to understand the predictive power of the respective inhibitor. Standard deviations and errors, and degrees of freedom (DF) were used as input for calculating the 95% probability interval. We have reported in the text the outcome of statistical tests in APA style, adapted to journal requirements. For relationships that appear non-linear – logarithmic or exponential – we have used the log10 function to transform the data if that makes the relationship appear linear, before re-applying linear regression. We have also used the log10 transformed datasets for the calculation of correlation coefficients, to correct for skewness.

Finally, we created a simple, discrete model resulting in one compound value, using selected flu-like inhibitors. This was to determine the optimal average threshold values for these inhibitors, which have the highest joint correlation with changes in flu-like incidence (ILI_3WMA_). We applied linear regression (F-test) to understand the predictive power of the compound value, and to determine the linear equation when significant. By constructing one compound, independent variable, we covered for collinearity or interaction effects between joined co-inhibitors. In our analysis, we based the compound value on three selected thresholds. For example, when one threshold value is passed, this leads to a compound value = 1, and when all three threshold values are passed, this leads to a compound value = 3. Therefore, the compound values are in the range of [0, 3].

The compound value equation can be expressed as shown below, where iv = the respective independent variable that acts as inhibitor of flu-like incidence, and k relates to the respective calculated threshold value. For each respective threshold passed (iv > k), +1 is added to the compound value (CV):

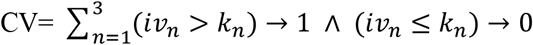

For the three selected co-inhibitors, this takes in Excel the form of CV = IF(IV_1_>K_1_;1;0) + IF(IV_2_>K_2_;1;0) + IF(IV_1_>K_3_;1;0), whereby we have used a threshold value for solar radiation (k_r_), and both pollen threshold values for allergenic (k_ap_) and total pollen (k_p_) for K_1_, K_2_ and K_3_ respectively, as is shown in section 3.

It is outside of the scope of this research to verify the underlying datasets of Elkerliek Ziekenhuis, RIVM/Nivel, CBS, and KNMI by examining the validity and reliability of their data collection methods. These institutes have well-established and internationally standardized protocols for data collection and verification.

All regression analyses have been carried out using the statistical package R version 3.5.

## 3. Results

The means and standard deviations per variable are given in Table 1. For the correlation coefficients below, we have used the log10 transformed data for respective variables, to correct for skewness.

**Table 1:** Overview of means (M) and standard deviations (SD) per variable in the dataset, including Log10 transformed data, and datasets that are reduced for sensitivity analysis.

| Variable | Mean | SD | Log10 transformed |  |
| --- | --- | --- | --- | --- |
|  |  |  | Mean | SD |
| Allergenic pollen concentration in grains/m3 | 349 | 987 | 1.84 | 0.86 |
| Total pollen concentration in grains/m3 | 732 | 1368 | 2.17 | 0.98 |
| Total pollen concentration in grains/m3 (minus weeks 42 to 50) | 870 | 1452 | 2.45 | 0.76 |
| Low-level allergenic pollen concentration in grains/m3 | 383 | 626 | 1.85 | 1.03 |
| Flu-like incidence (ILI) per 100K citizens | 47 | 40.2 | 1.54 | 0.35 |
| ΔILI | -0.25 | 15.4 | | |
| ΔILI<sub>3WMA</sub> | -0.26 | 8.9 | | |
| ΔILI<sub>3WMA</sub> (minus weeks 42 to 50) | -0.89 | 9.3 | | |
| Index hay fever | 101 | 116 | 1.81 | 0.39 |
| Relative humidity (%) | 79 | 8.27 |  |  |
| Temperature in °C | 10.8 | 5.82 |  |  |
| Solar radiation in J/cm2 | 1047 | 709 |  |  |
| Compound Model in thresholds passed | 1.4 | 1.10 |  |  |

When further inspecting the datasets regarding pollen concentrations and flu-like incidence reported by primary medical care in the Netherlands, it was clear that there are continuous pollen bursts (Figure 3), whereby only a few of these pollen bursts are classified as more allergenic (Figure 6). These pollen bursts, allergenic or low-level allergenic, typically coincide with and precede a decline in flu-like incidence.

**Figure 3:**
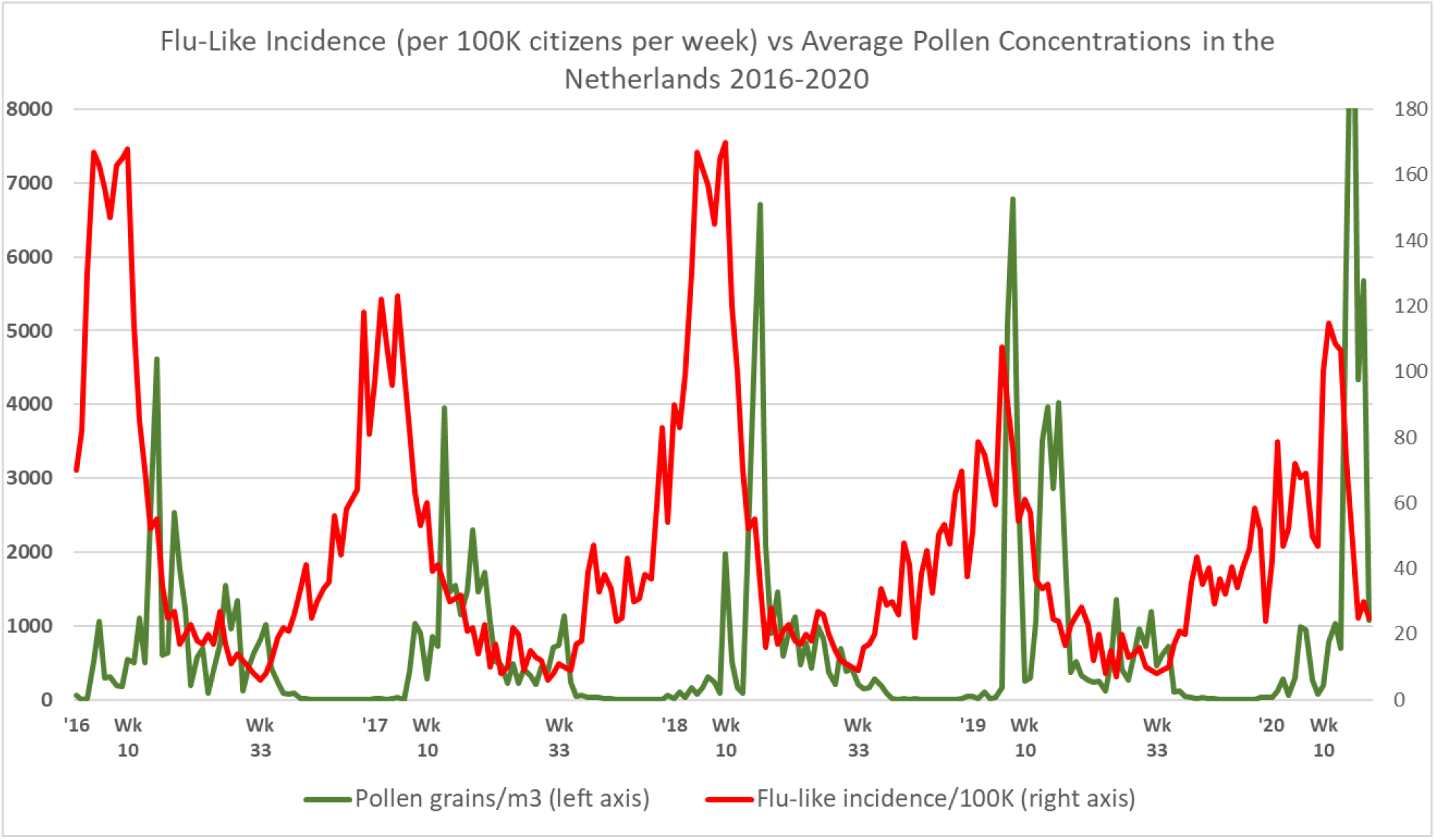
Total pollen concentrations vs. flu-like incidence in the Netherlands, whereby especially passing the 610 total pollen grains/m^3^ threshold marks the seasonal switches around week 10 (±5 weeks) and week 33 (±2 weeks). For the interpretation of the relationship, an incubation time of up to 2 weeks should be taken into account, and the change in flu-like incidence (Ro >1 or Ro <1) should be considered.

The correlation for total pollen and flu-like incidence is highly significant when taking into account incubation time: r(222) = −0.40, p < 0.001. We can thus reject the null-hypotheses H1_0_ in favor of the alternative hypothesis that, when taking into account incubation time, there is a negative correlation between total pollen and flu-like incidence, including the first cycle of the COVID-19 pandemic.

Furthermore, we can reject H5_0_ in favor of our assumption that it makes sense to also include low-level allergenic pollen concentrations in our study. Low-level allergenic pollen is inversely correlated to flu-like incidence (r(221) = −0.37, p < .00001), especially when corrected for the 2 weeks incubation time (r(219) = −0.53, p < .00001).

The fact that the correlations become stronger when taking into account incubation time, implies temporality. Furthermore, we can also observe from Figure 3 that flu-like incidence starts to decline *after* the first pollen bursts. Moreover, flu-like incidence starts to increase sharply *after* pollen concentrations become very low or close to zero. This is a qualitative indication of temporality. Furthermore, we can notice that the first COVID-19 cycle behaved according to pollen-flu seasonality, at least does not break with it.

When testing the impact on ΔILI, the weekly *changes* in medical flu-like incidence, the extended dataset till 2020, including COVID-19, shows a strong and highly significant inverse correlation with total pollen (r(222) = −0.26, p = 0.000089). Therefore, we can falsify the null-hypothesis (H2_0_) that there is no inverse correlation between the weekly pollen concentrations and weekly changes in flu-like incidence (ΔILI), including the period covering the first cycle of the COVID-19 pandemic. This inverse correlation therefore provides further support for the alternative hypothesis that the presence of an elevated level of pollen has an inhibiting effect on flu-like incidence, and starts to immediately influence the direction and course of the epidemic life cycle. Also, during the COVID-19 dominated period of the last 9 weeks, it appears that flu-like incidence behaves according to the expected pollen-flu seasonality. This strengthens the idea that COVID-19 might itself be seasonal, like all other flu-like pandemics since the end of the 19^th^ century. Also when studying other data from RIVM.nl about COVID-19 hospitalizations, we cannot conclude that COVID-19 breaks through the seasonal barrier. For example, new COVID-19 hospitalizations decreased from a peak of 611 on March 27 to just 33 on May 3, the last day of week 18.

Using the three-week moving average (ΔILI_3WMA_) of changes in flu-like incidence, the correlation coefficients become stronger and are again highly significant for total pollen concentration (r(223) = - 0.41, p < 0.00001). The bootstrapped correlation coefficient calculation gives a comparable outcome (r(223) = −0.38, p < 0.0001). As a second sensitivity analysis, we used the reduced dataset (minus the weeks of low pollen activity) and again found similar correlations (r(191) = −0.44, p < 0.0001; bootstrapped r(188) = −0.44, p < 0.0001, CI 95% −0.46 to −0.25)). We can thus also reject the null-hypothesis (H2_0_) that there is no inverse relationship between pollen and ΔILI_3WMA_, the three-week moving average of changes in flu-like incidence including incubation time. As this correlation (see also Figure 4) is stronger than it would be if it were not corrected for the incubation period, it is a further indication of temporality. And given that it is even stronger when the 2019/2020 flu-like season is included, it also does not contradict the idea that COVID-19 is subject to pollen induced flu-seasonality.

**Figure 4:**
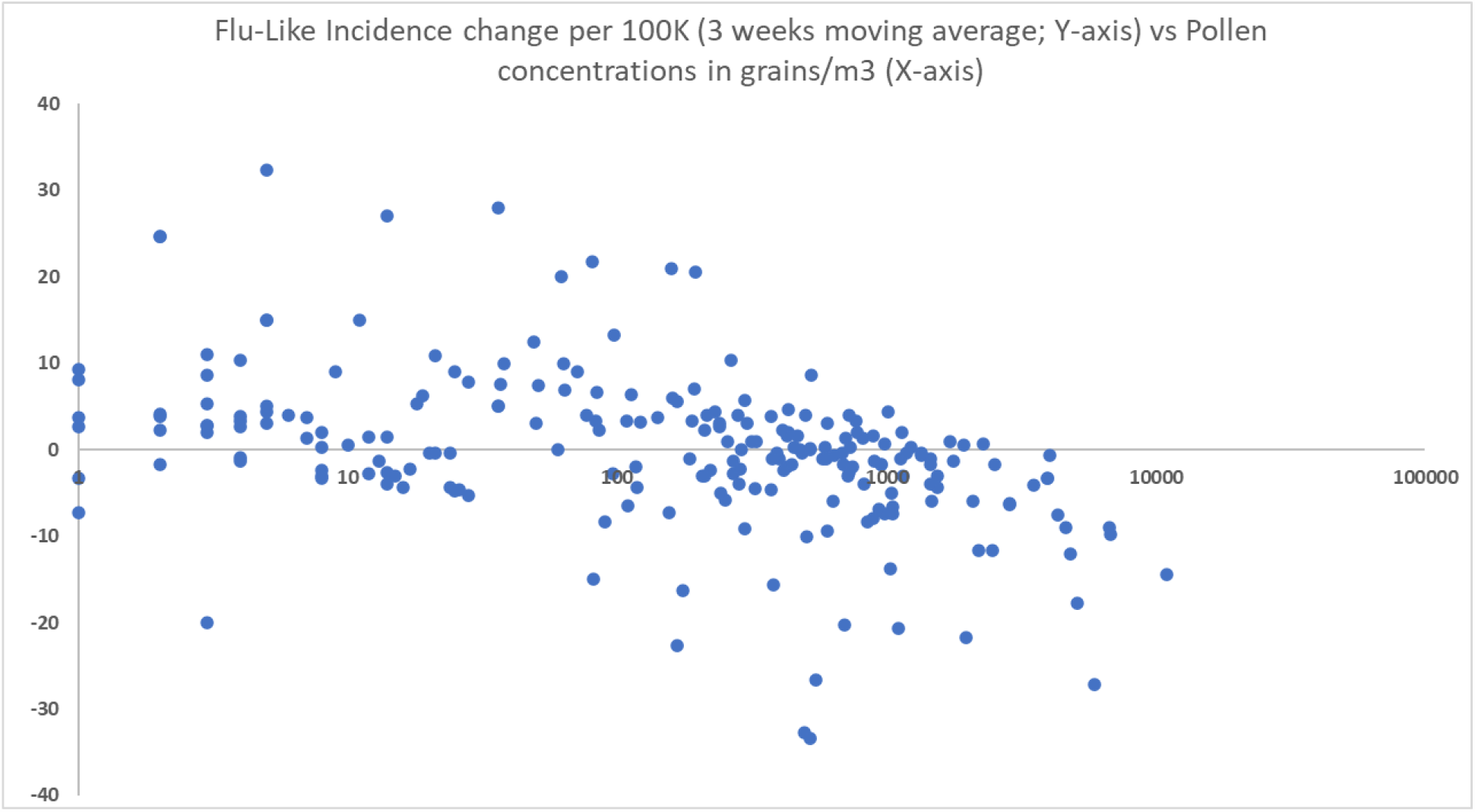
Scatter diagram showing the inverse relationship between changes in flu-like incidence (ILI_3WMA_) and log10(pollen concentrations in grains/m^3^).

Linear regression analysis shows that pollen has a highly significant inhibitory effect on flu-like incidence change (ΔILI_3WMA_) of F(1, 222) = 37.1, p < 0.001 (see Table 2, line 1), as a further basis for using total pollen concentration as a predictor. A Log10 transformation of pollen to compensate for visual non-linearity leads to a similar outcome: F(1, 219) = 43.87, p < 0.001 (see Table 2, line 4). At least visually, it is a good fit (see Figure 4).

**Table 2:**
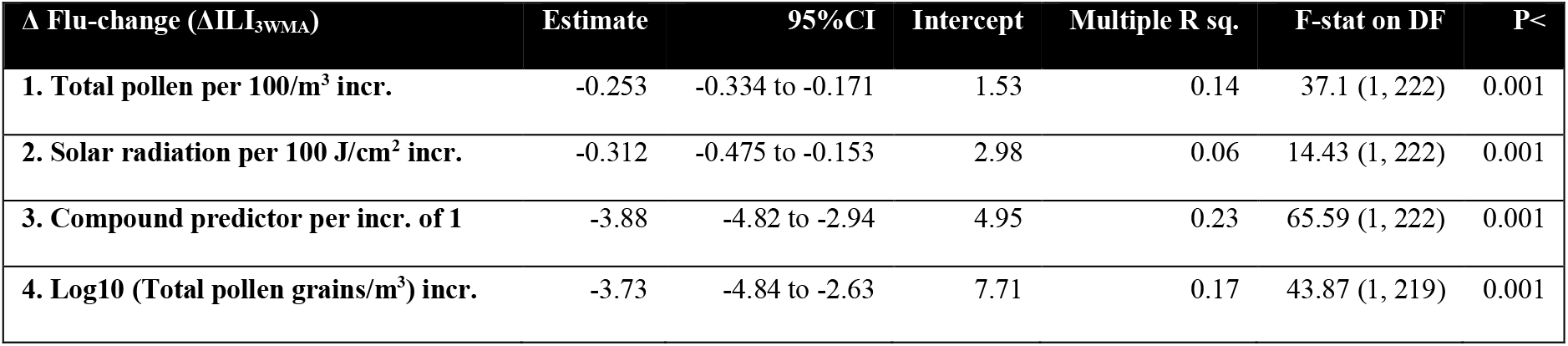
Summary of univariate regression analyses of pollen (1), solar radiation (2) and our compound pollen/solar radiation predictor (3) on changes in flu-like incidence (ΔILI_3WMA_). This shows all highly significant (p < 0.001) results, but the correlation for solar radiation is weak (0.06) and the compound predictor is the strongest (0.23). The log10 (4) analysis is a check on the non-linearity of pollen concentrations in relation to ΔILI_3WMA_, leading to the same conclusion.

In line with the correlation between pollen and flu-like incidence, the correlation between *total* pollen concentration and hay fever is stronger (r(162)= 0.76, p < 0.00001) than it is for allergenic and low-level allergenic pollen individually. This confirms that we can best use total pollen concentration as a predictor. Univariate regression analyses show that total pollen has a highly significant positive effect on hay fever incidence, which in turn has a highly significant inhibitory effect on flu-like incidence (see Table 3).

**Table 3:**
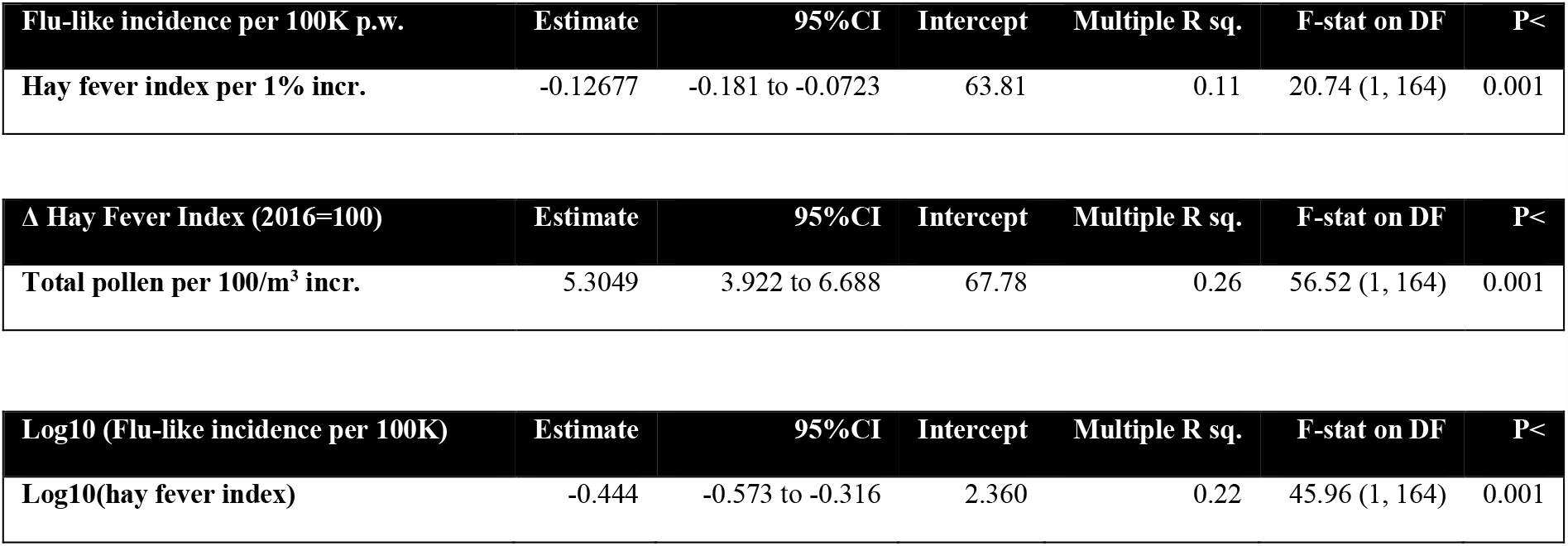
Summary of univariate regression analyses of total pollen concentration on hay fever (p < 0.001) and hay fever on flu-like incidence/100K citizens per week (p < 0.001), whereby pollen leads to an increase in hay fever, which in turn is associated with a decrease in flu-like incidence. To compensate for non-linearity, the regression of log10(hay fever) on log10(flu-like incidence) is added, with a similar, highly significant outcome.

Low-level allergenic pollen also has a highly significant effect on hay fever: r(160)= 0.77, p < 0.00001. We can thus reject the null-hypothesis H5_0_ in favor of the alternative hypothesis that low-level allergenic pollen also has a positive effect on hay fever. This might imply that pollen classified as none-to-low-level allergenic might still be responsible for certain allergic effects, and not just the more allergenic pollen. Therefore, trying to use low-level allergenic pollen to discriminate effects outside the allergenic path regarding the immune system might be challenging.

The nature of the relationship between hay fever and flu-like incidence might be statistically described as linear. However, it could be better described as logarithmic (Figure 5). In the context of this study, we have interpreted it as a further indication that it could also be described as a threshold-based switching pattern, conforming with the threshold-based approach that we have taken in our compound value, as calculated below.

**Figure 5:**
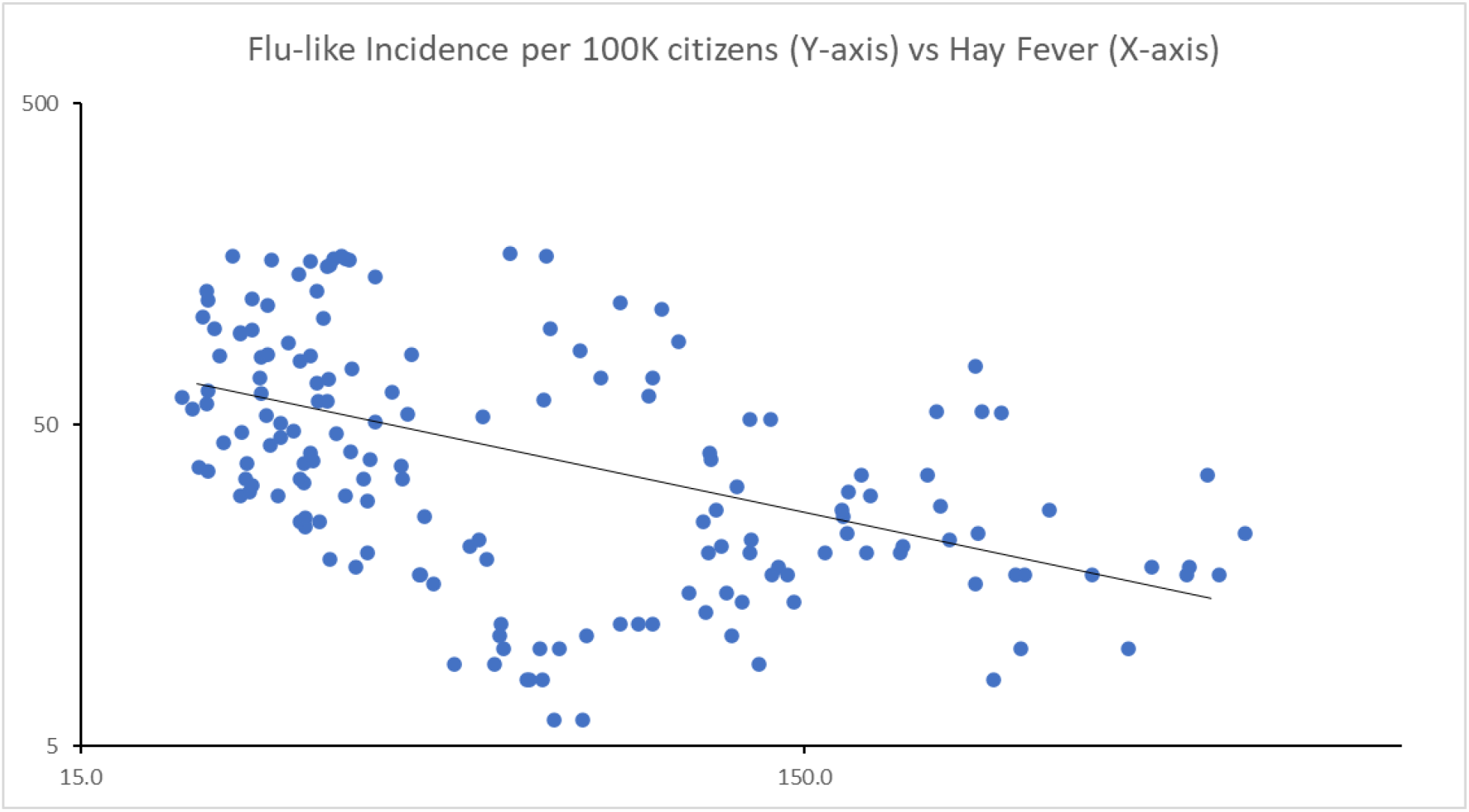
Inverse correlation between hay fever and flu-like incidence.

**Figure 6:**
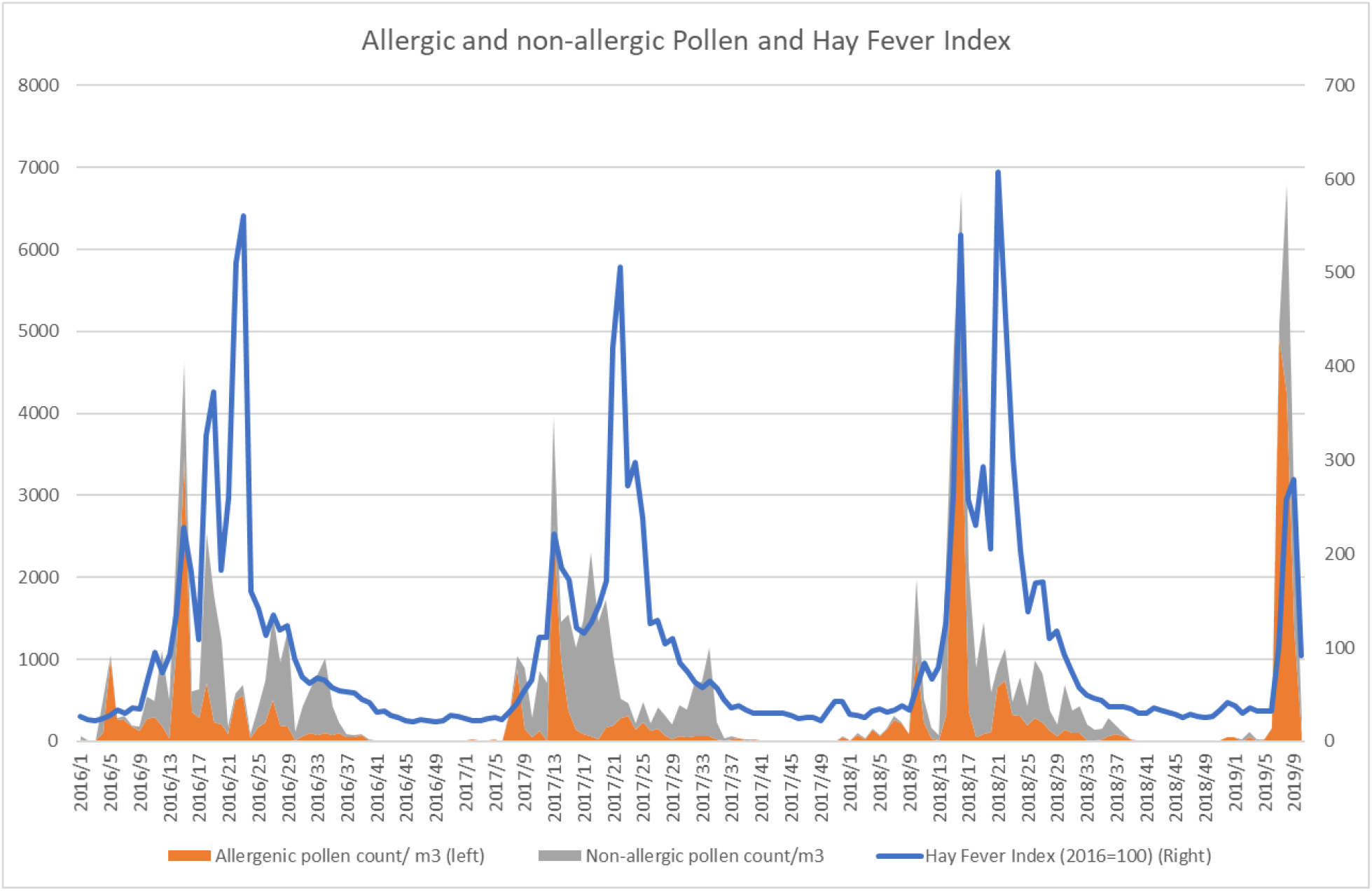
Both allergenic and low-level allergenic pollen are positively correlated to hay fever.

The expected effects of relative humidity (r(223) = −0.86, p < 0.0001), temperature (r(223) = 0.41, p < 0.0001) and solar radiation (r(223) = 0.67, p < 0.0001) on total pollen were found. So sunny, warmer and dry weather does indeed go hand-in-hand with an increase in pollen count. We can therefore reject the null-hypothesis that the selected meteorological variables (H4_0_) have no effect on pollen, whereby relative humidity reduces the amount of aerosol pollen.

Counter to findings in other studies, relative humidity is positively associated with changes in flu-like incidence (ΔILI_3WMA_) in the Netherlands (r(224) = 0.34, p < 0.00001). The Dutch flu season is cold and humid, and on rainy days the effect of pollen and solar radiation are reduced. Although temperature strongly correlates with flu-like incidence (r(226) = −0.82, p < 0.0001), it has a negligible effect on ΔILI, weekly changes in flu-like incidence (r(224) = −0.02 n.s.), also when corrected for incubation time. Therefore, it seems unlikely that temperature has a direct effect on aerosol flu-like viruses and the life cycle of a flu-like epidemic. In line with this, temperature is also not a good marker for the onset or the end of the flu season. In the Netherlands the end of the flu season (Ro<1) can coincide with an average temperature that is close to 0°C and the start of the flu season (Ro>1) can coincide with temperatures as high as 17°C.

Of the meteorological variables, only solar radiation has a highly significant inverse correlation with *changes* in flu-like incidence (ΔILI_3WMA_): (r(224) = −0.25, p = 0.000156).

Thus, of the meteorological variables, when it comes to solar radiation and relative humidity the null-hypothesis (H4_0_) can also be rejected, as they seem to effect the flu-like epidemic lifecycle. Of these two, only solar radiation is a flu-like inhibitor in line with its positive effect on pollen concentration, its association with immune-activation and the effect that UV has on viruses.

A univariate linear regression also shows the highly significant negative correlation for solar radiation on flu-like incidence change (ΔILI_3WMA_) (F(1, 222) = 14.43, p < 0.001 (see Table 2, line 2). As the correlation is weak (Multiple R-squared = .06), we have interpreted solar radiation as a co-inhibitor in relation to pollen; as a stand-alone independent variable its effect is too weak to explain flu-like seasonality.

Taking into account all these findings, we developed a discrete, compound model in which we included the changes in flu-like incidence (ΔILI_3WMA_), a threshold value for solar radiation (k_r_), and both pollen threshold values for allergenic (k_ap_) and total pollen (k_p_). We found that the compound model has the highest inverse correlation (r(222) = −0.48, p < 0.001) for the following threshold values: k_r_: 510 J/cm^2^, k_ap_: 120 allergenic pollen grains/m^3^, and k_p_: 610 total pollen grains/m^3^. The bootstrapped correlation coefficient calculation gives a comparable outcome (r(222) = −0.47, p < 0.0001). In line with the previous outcomes, the inclusion of relative humidity, low-level allergenic pollen or temperature did not improve the correlation strength of this model. Furthermore, given that they showed no significant interaction effects with pollen, it was not necessary to take such interactions into consideration in the model.

In each of the observed years, the now (re)defined pollen thresholds are passed in week 10 (± 5 weeks), depending on meteorological conditions controlling the pollen calendar and coinciding with reaching flu-like peaks, and again in week 33 (± 2 weeks), marking the start of the new flu-like season.

There is a highly significant inverse relationship between our compound threshold-based predictor value with flu-like incidence change (ΔILI_3WMA_) of F(1, 222) = 65.59, p < 0.001 and a Multiple R-squared correlation of 0.2281 (see Table 2, line 3). This confirms the usefulness of a discrete, pollen and solar radiation threshold-based model as a predictor of switches in flu-like seasonality, whereby the effect of pollen is stronger than that of solar radiation. As a consequence, we can reject the null-hypothesis (H3_0_) that this compound pollen/solar radiation value has no predictive significance for flu-like seasonality.

## 4. Discussion

First of all we will discuss the possible implications of the results for our theoretic model and alternative explanations. Next, we will discuss our methods.

### Theoretic model

We found highly significant inverse relationships between pollen and solar radiation and (changes in) flu-like incidence: a higher pollen concentration or an increase in solar radiation is related to a decline in flu-like incidence. This inverse correlation with pollen becomes stronger when the 2019/2020 period is included, which has been increasingly dominated by COVID-19 during the last 9 weeks. Given that more time will be needed to draw conclusions about whether COVID-19 is seasonal or not, from the data in this study it can only be observed that COVID-19 is not breaking with the flu-like seasonality pattern. Alternatively, social distancing may have contributed to flattening both the flu-like epidemic and COVID-19 pandemic curves at the tail-end of the 2019/2020 flu-like season. The Dutch government imposed hygiene measures from March 9, 2020 onwards and a mild form of a lockdown, that included social distancing, from March 11. Such behavioral policies will need to be included in the theoretic model, in addition to pollen and meteorological variables, if we want to be able to understand the relative importance of social distancing versus seasonality. We could, for example, more explicitly include behavioral variables (Gozzi et al, 2020) in the compound model, by rating lockdown regimes on a Likert-type scale [1, 5], from no lockdown (1) to a complete lockdown (5). Although seasonal behavior might be implicitly covered by the meteorological variables, it could still make sense to model them more explicitly as there might be cultural patterns in play – such as holidays or seasonal celebrations – that need to be taken into account.

The highly significant inverse correlation between hay fever and flu-like incidence confirms that allergic rhinitis makes it more difficult for flu-like viruses to propagate.

Solar radiation, the only meteorological variable that has a co-inhibitive effect on changes in flu-like incidence, has a stimulating effect on aerosol pollen formation and is responsible for melatonin-induced immuno-activation. Relative humidity reduces pollen aerosol formation, and correlates positively with flu-like incidence. We did not specifically look at precipitation, but it might make sense to explicitly consider this independent variable, given that it reduces pollen dissemination.

In our study we showed that temperature, aside from the fact that it influences pollen, has no predictive value for changes in flu-like incidence. Therefore, its inverse correlation with flu-like incidence might be interpreted in a number of ways: a) as spurious: the common causal factor is solar radiation, or b) as a stressor that has immediate effects on the functioning of the immune system of already infected persons. When discussing the influence of meteorological variables, we assume that the associated behavioral aspects are covered. These are sometimes summarized as seasonal behavior, but this independent variable might have a cultural dimension that needs to be better understood.

We showed that a compound value, based on threshold values for pollen and solar radiation, results in a stronger correlation with the flu-like lifecycle than the individual inhibitors. This model could form an empirical basis for understanding flu-like seasonality, its Ro and reliably predicting the start and end of each flu-like cycle. Given that behavior, in the form of hygiene and social distancing, is also widely seen as an inhibitor, it might be worthwhile to also include this factor in our compound value. This will probably lead to an even stronger predictor for the evolution of the reproduction number Ro of flu-like epidemics, although this might be beyond explaining the seasonality effect itself.

For as long as the level of herd immunity (Fine et al., 2011) for COVID-19 is still below required thresholds for ending pandemics (Plans-Rubio, 2012), it might make sense to also include indications of herd immunity levels in the theoretic model.

Finally, despite pollution not been seen as an inhibitor of flu-like incidence (Coccia, 2020), it still might interact with pollen. A more complete theoretic model, controlling for the (interactions with) pollution, could give more insight in how to interpret the findings of this or similar studies.

### Methodological considerations

In general, statistical research cannot prove causal relationships in uncontrolled environments, even if datasets seem to behave *as if* there is causality. Such statistics, however, can provide indications and identify reliable predictors, help filter out bad ideas, and be the inspiration for testable hypotheses that can be verified in laboratory and other fully controlled experiments.

Although the datasets seem to be sufficiently representative, there appears to be room for improvement. For example, including the data of more weather stations might help to improve the approximation of the weather conditions the Dutch population experiences on average, and help to distinguish patterns per province. Furthermore, it might be useful to include wind speeds, given that these constitute a vector for the dispersal of pollen in the Netherlands, which has a maritime and temperate climate. Additionally, the effects of climate change on pollen maturation (Frei & Gassner, 2008) might also be an important factor. Another example of improving the representativeness would be by including more pollen types in the particle counts than are currently covered by the current methodology of the European Allergy Network.

Specifically, the validity and reliability of the hay fever index is unclear, and the dataset is not maintained. There might be lag effects, but these are unknown. It might also be a good idea to include alternative datasets, such as search-engine based trend analysis to be able to generate a complete dataset for a whole period of study. These could be validated separately. Alternatively, representative medical datasets could be obtained.

Finally, the ILI dataset, which is now based on a sample of 40 representative local primary care units, could be improved. For example, including *all* patients visiting *any* primary care unit in the Netherlands would reduce the need for extrapolation, with its inherent risk of bias. The use of Δ ILI_3WMA_ including incubation time, seems to be a more elegant option than carrying out multiple tests with ΔILI. At the same time, it could be argued that this metric might be better as a two-week forward-looking moving average. This is because it is unlikely that any effects will be noticed in the first week, given the average reporting delays and incubation period.

## 5. Conclusion

We conclude that pollen and solar radiation both have highly significant inverse correlations with changes in flu-like incidence. Furthermore, the inverse correlation between pollen and (changes in) flu-like incidence becomes stronger when the incubation time is included. A compound variable – based on the thresholds for total pollen, allergenic pollen and solar radiation – shows the strongest correlation with changes in flu-like incidence, and appears to be useful as a predictor for switches in flu-like seasonality.

COVID-19 has dominated the tail-end of the 2019-2020 flu-like season in the Netherlands. And, although COVID-19 *appears* to be also subject to flu-like seasonality, like pandemics that preceded it, it is still too early to draw conclusions about this.

It will require further research to test the findings, threshold values and predictive model for flu-like seasonality in other countries with different climates. Controlled experiments are needed to deepen the biomedical understanding of how allergic rhinitis and immuno-activation by pollen protects against the spread of flu-like viruses, and to confirm and understand the assumed interaction between pollen and viral bio-aerosol.

## Data Availability

All datasets are based on public datasets in The Netherlands to which we have referred in the paper, and our datasets are available for review.

## Acknowledgements

thanks to Sowjanya Putrevu, data scientist at Icecat, for her voluntary support with<u>in</u> executing statistical tests.

